# *NPC1* variants are not associated with Parkinson’s Disease, REM-sleep behaviour disorder or Dementia with Lewy bodies in European cohorts

**DOI:** 10.1101/2022.11.08.22281508

**Authors:** Emma N. Somerville, Lynne Krohn, Eric Yu, Uladzislau Rudakou, Konstantin Senkevich, Jennifer A. Ruskey, Farnaz Asayesh, Jamil Ahmad, Dan Spiegelman, Yves Dauvilliers, Isabelle Arnulf, Michele T.M. Hu, Jacques Y. Montplaisir, Jean-François Gagnon, Alex Desautels, Abubaker Ibrahim, Ambra Stefani, Birgit Hogl, Gian Luigi Gigli, Mariarosaria Valente, Francesco Janes, Andrea Bernardini, Petr Dusek, Karel Sonka, David Kemlink, Giuseppe Plazzi, Elena Antelmi, Francesco Biscarini, Brit Mollenhauer, Claudia Trenkwalder, Friederike Sixel-Doring, Michela Figorilli, Monica Puligheddu, Valerie Cochen De Cock, Luigi Ferini-Strambi, Anna Heibreder, Christelle Charley Monaca, Beatriz Abril, Femke Dijkstra, Mineke Viaene, Bradley F. Boeve, Ronald B. Postuma, Guy A. Rouleau, Ziv Gan-Or

## Abstract

*NPC1* encodes a lysosomal protein involved in cholesterol transport. Biallelic mutations in this gene may lead to Nieman-Pick disease type C, a lysosomal storage disorder. The role of *NPC1* in alpha synucleinopathies is still unclear, as different genetic, clinical, and pathological studies have reported contradictory results. This study aimed to evaluate the association of *NPC1* variants with the synucleinopathies Parkinson’s disease (PD), dementia with Lewy bodies (DLB), and rapid eye movement (REM)-sleep behavior disorder (RBD). We analyzed common and rare variants from three cohorts of European descent: 1,084 RBD cases and 2,945 controls, 2,852 PD cases and 1,686 controls, and 2,610 DLB cases and 1,920 controls. Logistic regression models were used to assess common variants while optimal sequence Kernel association tests (SKAT-O) were used to assess rare variants, both adjusted for sex, age, and principal components. No variants were associated with any of the synucleinopathies, supporting that common and rare *NPC1* variants do not play an important role in alpha synucleinopathies.

## 1. Introduction

Alpha synucleinopathies are a group of neurodegenerative disorders characterized by alpha-synuclein and Lewy body accumulation in the brain. These disorders include mainly Parkinson’s disease (PD), dementia with Lewy bodies (DLB), multiple system atrophy (MSA), and their prodromal rapid-eye-movement (REM)-sleep behavior disorder (RBD). Alpha-synucleinopathies have complex etiology and may show overlap with other disorders. One such group of disorders is lysosomal storage disorders (LSDs), characterized by a lysosomal accumulation of undegraded substrates, often due to the dysfunction of various lysosomal enzymes (Filocamo and Morrone, 2011). Many commonalities have been observed in both groups, including alpha-synuclein deposits in the brain, substantia nigra pathology, lysosomal dysfunction, lipid metabolism alterations, and impairment of the ubiquitin-proteasome system (Shachar et al., 2011). Additionally, a number of genes, including *GBA, SMPD1, ASAH1* and *GALC*, have been identified as contributors to both alpha-synucleinopathies and LSDs (Alcalay et al., 2019; Deane et al., 2011; Nalls et al., 2019; Riboldi and Di Fonzo, 2019; Robak et al., 2017).

Biallelic mutations in the sphingomyelin phosphodiesterase 1 (*SMPD1)* gene lead to Niemann-Pick disease Types A and B, and heterozygous *SMPD1* variants have been associated with PD and DLB (Alcalay et al., 2019; Clark et al., 2015; Desnick et al., 2010). The third class of Niemann-Pick disease, Type C (NPC), is caused by variants in the NPC intracellular cholesterol transporter 1 gene (*NPC1*). NPC has also been connected to synucleinopathies, as multiple *NPC1* variant carriers have been reported to display PD symptoms and Lewy body deposits in the brain (Chiba et al., 2013; Josephs et al., 2004; Kluenemann et al., 2013; Schneider et al, 2019). A genetic study has attempted to examine the relationship between *NPC1* variants and PD, finding no association (Ouled Amar Bencheikh et al., 2020). However, this study may have been underpowered and did not examine associations with other synucleinopathies.

In the present study we aimed to further elucidate the role of *NPC1* in alpha-synucleinopathies, by testing if common and rare variants in *NPC1* were associated with PD, DLB and RBD in a total of 6,546 patients and 6,551 controls.

## 2. Methods

### 2.1 Study Population

The study population consisted of three separate cohorts: 1) an RBD cohort of 1,084 cases and 2,945 controls collected through the International RBD Study Group (IRBDSG) at McGill University, 2) a PD cohort consisting of 2,852 cases and 1,686 controls obtained from the Accelerated Medicines Partnership: Parkinson’s Disease (AMP-PD, https://amp-pd.org/) program, and 3) a DLB cohort including 2610 cases and 1920 controls obtained from dbGaP (phs001963.v1.p1). AMP-PD patient data included the Parkinson’s Progression Markers Initiative (PPMI), the Parkinson’s Disease Biomarkers Program (PDBP), the Harvard Biomarker Study (HBS), the STEAD-III Investigators study, and the Fox Investigation for New Discovery of Biomarkers (BioFIND). AMP-PD data for controls came from PPMI, PDBP, HBS, BioFIND and the International LBD Genomics Consortium (iLBDGC). Demographics for the three cohorts can be found in Table 1. Due to a lack of sequencing data for other populations only subjects of European descent were included, determined through principal component analysis. RBD was diagnosed with video-polysomnography according to the International Classification of Sleep Disorders (ICSD) version 2 or 3 (American Academy of Sleep Medicine, 2014; American Academy of Sleep Medicine, 2005; Hogl and Stefani, 2017). Informed consent forms were signed by all participants before entering the studies and study protocols were approved by the institutional review boards.

**Table 1.**
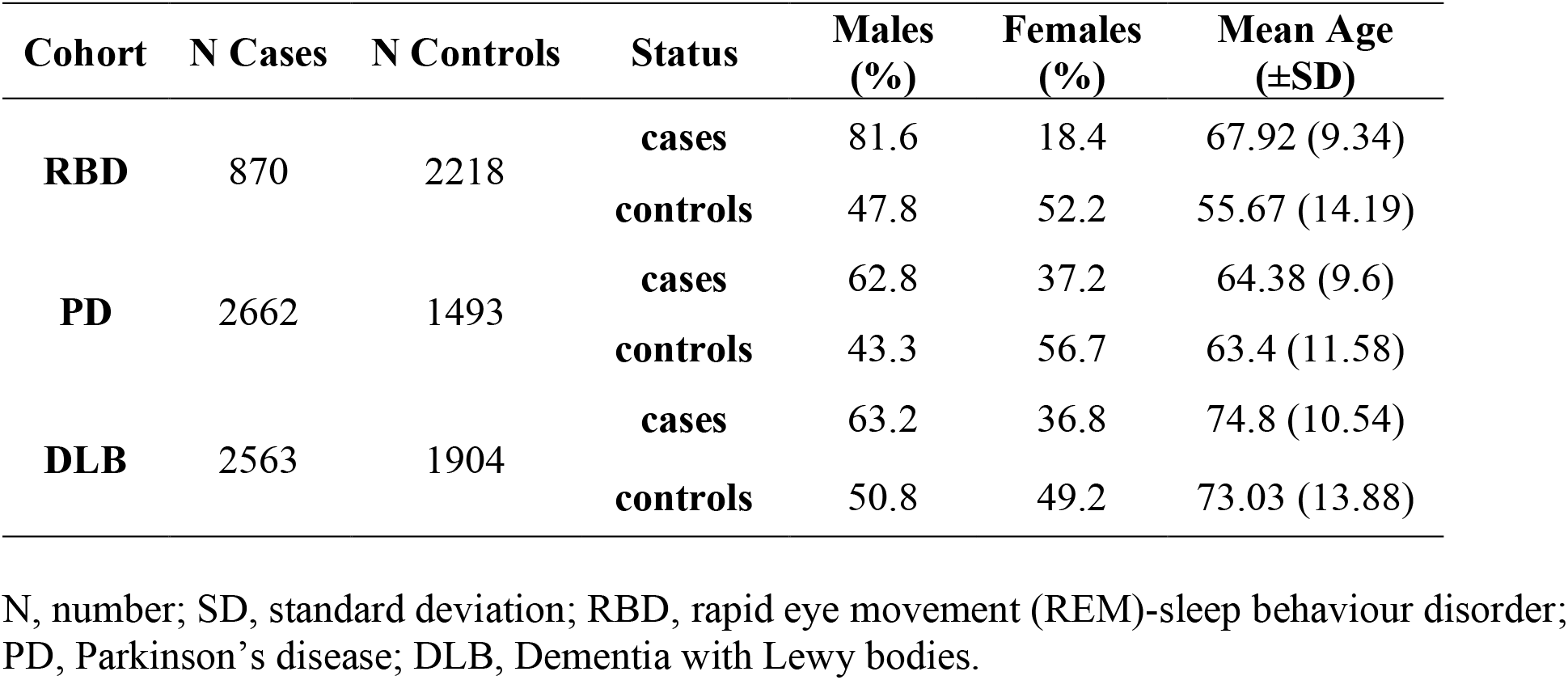
Cohort demographics and individuals analyzed after quality control.

### 2.2 NPC1 Sequencing

We used molecular inversion probes (MIPs) to target the coding and regulatory sequence of *NPC1* in the RBD cohort, as previously described (O’Roak et al., 2012). MIPs used to sequence *NPC1*, as well as the targeted DNA capture and amplification method, have also been described (Ross et al., 2016; Rudakou et al., 2020). Sequencing was performed using Illumina HiSeq 2500/4000 and NovaSeq 6000 at the Genome Quebec Innovation Centre. Reads were then mapped to the human reference genome (hg19) using Burrows-Wheeler Aligner (Durbin, 2009). Post-alignment quality control and variant calling was performed using Genome Analysis Toolkit (GATK, v3.8) as previously described (McKenna et al., 2010), and the Ensembl Variant Effect Predictor (VEP) was used to annotate variants (McLaren et al., 2016).

Whole genome sequencing (WGS) data and clinical data were obtained from dbGaP (phs001963.v1.p1) for the DLB cohort and AMP-PD https://amp-pd.org/whole-genome-data) for the PD cohort. The NPC1 gene region was extracted from these datasets using PLINK v1.9 (Purcell et al., 2007).

### 2.3 Quality Control

Quality control for the RBD cohort (MIPs data) was performed using PLINK v1.9 (Purcell et al., 2007) and Genome Analysis Toolkit (GATK) as previously described (Ouled Amar Bencheikh et al., 2020). In brief, variants were excluded if coverage depth was lower than 50x and if the variant appeared in less than 25% of the reads. We further excluded variants based on the following criteria: genotyping rate less than 90%, missingness difference between cases and controls set at *p* = 0.05 after Bonferroni correction, and deviation from the Hardy Weinberg equilibrium set at *p* = 0.001. Samples were removed if their average genotyping rate was less than 90%. The pipeline is available at https://github.com/gan-orlab/MIPVar.

Sequence data for the DLB and PD cohorts was run through standard genome-wide association study (GWAS) quality control for samples and variants as previously described (https://github.com/neurogenetics/GWAS-pipeline) (Nalls et al., 2019).

### 2.4 Statistical Analyses

To test for associations between common variants (MAF > 0.01) and rare variants (MAF < 0.01) in *NPC1* and disease status, we performed logistic regression for all three cohorts using PLINK v1.9 (Purcell et al., 2007). Optimized sequence Kernel association tests (SKAT-O, R program) were also performed on rare variants for each cohort, including tests for all rare variants, nonsynonymous variants, and pathogenic variants, as well as a combination of nonsynonymous, frameshift, splice and stop-gain variants (Lee et al., 2012). Pathogenic variants were determined as those with predominantly pathogenic or likely pathogenic reports in the ClinVar database, extracted using VEP (McLaren et al., 2016). All regression and SKAT-O analyses across the three cohorts included adjustment for sex, age and the top 10 principal components (based on scree plots).

## 3. Results

The average coverage of *NPC1* sequencing reads was 903x. We included reads with coverage above 50x, which comprised 96% of all *NPC1* reads. The number of cases and controls after quality control for each cohort can be found in Table 1. There were 4, 94 and 120 common *NPC1* variants (Supplementary Table 1) tested for association with RBD, PD and DLB, respectively. There were 57, 324 and 1924 rare *NPC1* variants, tested for association with RBD, PD and DLB (Supplementary Table 2). None of the individual common or rare variants were associated with any of the disease phenotypes after multiple testing correction (Supplementary Table 1 and 2). Additionally, no variant categories in the SKAT-O analyses were associated (Table 2).

**Table 2.** SKAT-O analyses of rare variants (MAF < 0.01)

| Variant category | RBD |  | PD |  | DLB |  |
| --- | --- | --- | --- | --- | --- | --- |
|  | N Variants | P-value | N Variants | P-value | N Variants | P-value |
| **All Rare** | 57 | 0.6654 | 324 | 0.3024 | 1924 | 0.4033 |
| **NS** | 14 | 0.5801 | 25 | 0.6189 | 67 | 0.2553 |
| **NS + FS + SG + splice** | 17 | 0.2957 | 29 | 0.445 | 78 | 0.4637 |
| **Pathogenic + FS** | 3 | 0.5298 | 7 | 0.5198 | 12 | 0.2955 |

SKAT-O, optimized sequence Kernel association test; RBD, rapid eye movement (REM)-sleep behaviour disorder; PD, Parkinson's disease; DLB, Dementia with Lewy bodies; N, number; NS, non-synonymous; FS, frameshift; SG, stop-gain.

In total, we found 17 variants identified predominantly as pathogenic or likely pathogenic for *NPC1* across our three cohorts. All of these variants were rare, with none showing association individually or when grouped together in the SKAT-O analyses (Supplementary Table 2).

We also investigated variants linked to the appearance of PD-like symptoms in NPC patients and *NPC1* variant carriers (Chiba et al., 2013; Josephs et al., 2004; Kluenemann et al., 2013; Schneider et al, 2019). A total of 75 cases and 57 controls were carriers of p.N222S (rs55680026) across all cohorts, with the variant being more common in controls in RBD and more common in cases in PD and DLB. The variant p.I1061T (rs80358259) was carried by 6 cases and 2 controls, being found more commonly in cases in PD and only in cases in DLB. There were 6 cases and 6 controls with p.S1004L (rs150334966), which was more common in cases than controls in PD and more common in controls than cases in DLB. Variants p.G992R and p.P1007A (rs80358254 and rs80358257) were found in the DLB cohort as well, but only in 1 control and 2 cases, respectively. Despite many of these variants being more frequent in cases, none were found to be significantly associated within any cohort. Further carrier details and association statistics can be found in Supplementary Table 2.

## 4. Discussion

In the present study, we performed thorough genetic association testing of *NPC1* in three cohorts of alpha synucleinopathies - RBD, PD and DLB. We did not find any association between *NPC1* variants and these disorders, supporting that *NPC1* does not play a major role in alpha synucleinopathies. Our results also support the previously mentioned study on *NPC1* and PD, which included an independent PD cohort and found no association of common and rare variants with disease risk (Ouled Amar Bencheikh et al., 2020).

Several cases of individuals who carried *NPC1* variants and had PD symptoms and Lewy bodies have been reported (Chiba et al., 2013; Josephs et al., 2004; Kluenemann et al., 2013; Schneider et al, 2019). However, none of the variants in our cohorts were associated with any synucleinopathy. Our findings, together with previous studies (Ouled Amar Bencheikh et al., 2020, Zech et al., 2013), suggest that heterozygous *NPC1* variants are unlikely to be an important risk factor for PD symptoms.

The lack of association of *NPC1* variants with alpha synucleinopathies, despite the demonstrated association of *SMPD1* with PD and DLB, could be due to biological pathway differences (Alcalay et al., 2019; Clark et al., 2015). *SMPD1* produces a lysosomal enzyme that breaks down sphingomyelin in the sphingolipid metabolism pathway (Rudakou et al., 2020).

Many genes that have been firmly shown to associate with synucleinopathies are in the same pathway, including *GBA, GALC, SMPD1*, and *ASAH1. NPC1*, however, is not in this pathway, and instead is responsible for the trafficking of cholesterol and lipids into the lysosome (Bosch and Kielian, 2015). Thus, the lack of association of *NPC1* with synucleinopathies supports that specific dysfunction of the sphingolipid metabolism pathway may be the key lysosomal disease mechanism in synucleinopathies.

This study has a few key limitations. First, we only included individuals of European ancestry, as there was no sufficient available genetic data from non-European populations to perform these analyses. The synucleinopathies included in the present study are complex disorders, thus it is possible that *NPC1* variants may contribute to RBD, DLB or PD in other populations. Another limitation is that there are differences in age and sex between patients and controls across all three cohorts. Both common and rare variant analyses were adjusted for sex and age to account for these differences. Lastly, our study cannot rule out the role of very rare *NPC1* variants in alpha-synucleinopathies, or variants not present in our cohorts. Rare variants present in only a few cases or controls, such as p.I1061T, p.G992R, and p.P1007A, should be inspected in a larger cohort to confidently determine their lack of association.

In conclusion, our results do not support a major role for *NPC1* in RBD, PD and DLB. However, larger studies will be required to definitively rule out *NPC1* as a contributing factor.

## Data Availability

REM-sleep behaviour disorder data produced in the present study is available upon request to the authors. Summary Statistics are available in the supplementary tables. Parkinson's disease data used in the present study is available online at https://amp-pd.org/. Dementia with Lewy bodies data used in the present study is available online at https://www.ncbi.nlm.nih.gov/projects/gap/cgi-bin/study.cgi?study_id=phs001963.v1.p1.
All results produced in the present study are contained in the manuscript.

## 5. Acknowledgements

We thank the participants for contributing to this study. We thank Meron Teferra, Helene Catoire and Sandra B. Laurent for their contributions. The work performed in this study was financially supported by grants from the Michael J. Fox Foundation, the Canadian Consortium on Neurodegeneration in Aging (CCNA), the Canadian Institutes of Health Research (CIHR), the Canada First Research Excellence Fund (CFREF), awarded to McGill University for the Healthy Brains for Healthy Lives initiative (HBHL), and Parkinson Canada. J-FG holds a Canada Research Chair on Cognitive Decline in Pathological Aging.

DLB data used in preparation for this article were obtained from dbGaP. More information on this study can be found at phs001963.v1.p1, and we acknowledge principal investigators Bryan J. Traynor, MD, PhD, and Sonja W. Scholz, MD, PhD for their work on producing the data. This study was funded by the National Institutes of Health.

PD data used in the preparation of this article were obtained from the AMP PD Knowledge Platform. For up-to-date information on the study, https://www.amp-pd.org. AMP PD – a public-private partnership – is managed by the FNIH and funded by Celgene, GSK, the Michael J. Fox Foundation for Parkinson’s Research, the National Institute of Neurological Disorders and Stroke, Pfizer, and Verily. ACCELERATING MEDICINES PARTNERSHIP and AMP are registered service marks of the U.S. Department of Health and Human Services.

## 6. Conflicts of interest

ZGO received consultancy fees from Lysosomal Therapeutics Inc. (LTI), Idorsia, Prevail Therapeutics, Inceptions Sciences (now Ventus), Ono Therapeutics, Denali, Handl Therapeutics, Neuron23, Bial Biotech, UCB, Guidepoint, Lighthouse and Deerfield.

## Notes

### Competing Interest Statement

Ziv Gan-Or has received consultancy fees from Lysosomal Therapeutics Inc. (LTI), Idorsia, Prevail Therapeutics, Inceptions Sciences (now Ventus), Ono Therapeutics, Denali, Handl Therapeutics, Neuron23, Bial Biotech, UCB, Guidepoint, Lighthouse and Deerfield.

### Clinical Protocols

https://github.com/gan-orlab/MIPVar

### Author Declarations

Ethics committee/IRB of McGill University gave ethical approval for this work

